# SARS-CoV-2 Omicron neutralization and risk of infection among elderly after a booster dose of Pfizer vaccine

**DOI:** 10.1101/2022.03.30.22273175

**Authors:** Timothée Bruel, Laurie Pinaud, Laura Tondeur, Delphine Planas, Isabelle Staropoli, Françoise Porrot, Florence Guivel-Benhassine, Mikaël Attia, Stéphane Pelleau, Tom Woudenberg, Cécile Duru, Aymar Davy Koffi, Sandrine Castelain, Sandrine Fernandes-Pellerin, Nathalie Jolly, Louise Perrin De Facci, Emmanuel Roux, Marie-Noëlle Ungeheuer, Sylvie Van Der Werf, Michael White, Olivier Schwartz, Arnaud Fontanet

**Affiliations:** Virus & Immunity Unit, Institut Pasteur, Université Paris Cité, CNRS UMR 3569, Paris, France; Vaccine Research Institute, Créteil, France; Emerging Diseases Epidemiology Unit, Institut Pasteur, Université Paris Cité, Paris, France; Molecular Genetics of RNA Viruses Unit, Institut Pasteur, Université Paris Cité, Paris, France; Infectious Disease Epidemiology and Analytics Unit, Institut Pasteur, Université Paris Cité, Paris, France; Hôpital de Crépy-en-Valois, Crépy-en-Valois, France; Laboratoire de virologie, CHU Amiens, AGIR UR4294, UPJV, Amiens, France; Center for Translational Science, Institut Pasteur, Paris, France; Clinical Investigation and access to bioresources (ICAReB) platform, Center for Translational Science, Institut Pasteur, Paris, France; Conservatoire National des Arts et Métiers, PACRI Unit, Paris, France

## Abstract

**Background:** The protective immunity against Omicron following a BNT162b2 Pfizer booster dose among elderly is not well characterized.

**Methods:** Thirty-eight residents from three nursing homes were recruited for the study. Antibodies targeting the Spike protein of SARS-CoV-2 were measured with the S-Flow assay. Neutralizing activities in sera were measured as effective dilution 50% (ED50) with the S-Fuse assay using authentic isolates of Delta and Omicron.

**Results:** Among the 38 elderly included in the study, with median (inter-quartile range, IQR) age of 88 (81-92) years, 30 (78.9%) had been previously infected. The ED50 of neutralization were lower against Omicron than Delta, and higher among convalescent compared to naive residents. During an Omicron epidemic affecting two of the three nursing homes in December 2021-January 2022, 75% (6/8) of naive residents got infected, compared to 25% (6/24) of convalescents (P=0.03). Antibody levels to Spike and ED50 of neutralization against Omicron after the BNT162b2 booster dose were lower in those with breakthrough infection (n=12) compared to those without (n=20): median of 1256 vs 2523 BAU/mL (P=0.02) and median ED50 of 234 vs 1298 (P=0.0004), respectively.

**Conclusion:** This study confirmed the importance of receiving at least three antigenic exposures to the SARS-CoV-2 Spike protein for achieving satisfactory neutralizing antibody levels. In this population, protection against Omicron infection was increased in individuals who had been previously infected in addition to the three vaccine doses. Thus, a fourth antigenic exposure may be useful in the elderly population to prevent infection with Omicron, a variant known for its high escape immunity properties.

## Introduction

The neutralization capacity of sera from vaccinated or convalescent individuals against the Omicron (B.1.1.529) (BA.1) variant of Severe Acute Respiratory Coronavirus 2 (SARS-CoV-2) has been well studied among several population groups, and has been shown to be lower against Omicron compared to other variants [1–12]. Information on the vaccine efficacy and the neutralization capacity of sera from elderly against Omicron is more limited [13,14], despite decreased immunogenicity, increased risk of severe forms of disease and accelerated waning of immunity in this population [15–17]. Here, we evaluated the capacity of a booster dose of BNT162b2 to elicit neutralizing antibodies against Omicron and examined levels of humoral immunity before Omicron breakthrough infections among residents living in nursing homes.

## Methods

Thirty-eight residents from three nursing homes were recruited from the Covid-Oise study (NCT04644159). This community-based cohort started in November 2020 and aims to monitor the immunological response following SARS-CoV-2 infection and Coronavirus Disease 2019 (COVID-19) vaccination.

Past infection of the residents was determined based on clinical data and detection of SARS-CoV-2 specific antibodies using two serological assays, as previously described [18,19]. All sera samples available since inclusion of the residents in the study were evaluated with both assays. Detection of past infection relied on anti-Spike (S) and anti-Nucleocapsid (N) antibodies for pre-vaccination sera, and only on anti-N antibodies for post-vaccination sera.

Sera were obtained two months (median 56 days, inter-quartile range [IQR] 52-61 days) after the second dose, and two months (median 55 days, IQR 49-59 days) after the third dose of mRNA vaccine (Figure 1a). All residents have been immunized with BNT162b2 for their primary series except four who received mRNA-1273. The two initial doses were received three weeks apart (median 21 days, IQR 21-21 days). All residents received a dose of BNT162b2 as booster eight months after the second dose (median 236 days, IQR 230-237 days).

**Figure 1:**
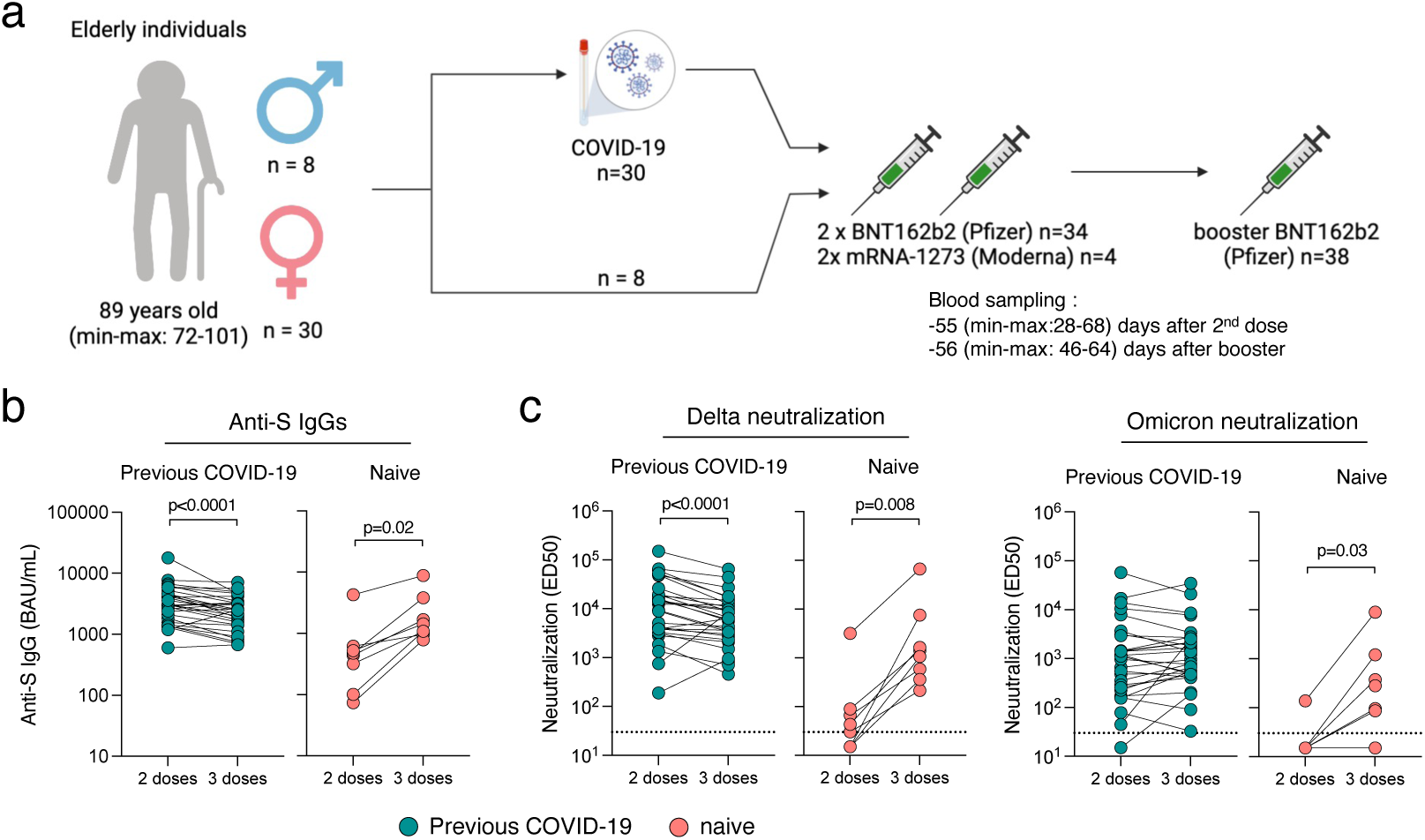
Immunogenicity of a booster dose of BNT162b2 vaccine in elderly individuals. (a) Thirty-eight elderly individuals from three households, (30 females and 8 males) were included in the analysis. All received a two-dose regimen of mRNA vaccine (Pfizer BNT162b2; n=34 or Moderna; n=4) and a booster dose (Pfizer BNT162b2; n=38) 8 months apart. Thirty were diagnosed with COVID-19 prior to their booster dose. (b) Anti-Spike IgGs were measured using the S-Flow assay 2 months after the second dose and 2 months after the booster dose. Data are provided as Binding Arbitrary Unit per mL (BAU/mL), the standardized WHO unit. The dashed line indicates the limit of detection. Comparisons were performed using the Wilcoxon matched-pairs signed rank test. (c) Neutralization of Delta and Omicron were measured using the live-virus assay S-Fuse 2 months after the second dose and 2 months after the booster dose. Data are provided as Effective Dilution 50 (ED50), indicating the dilution of serum capable of inhibiting 50% of viral infection. Green dots indicate individuals with an history of COVID-19 prior to their booster dose of vaccine. Pink dots indicate individuals with no previous COVID-19. The dashed line indicates the limit of detection. Comparisons were performed using the Wilcoxon matched-pairs signed rank test.

Antibodies targeting the Spike protein of SARS-CoV-2 were measured with the S-Flow assay (N=75). Briefly, this assay uses 293T cells stably expressing the Spike (S) protein (293T-S cells) and 293T control cells as control to detect anti-Spike antibodies by flow cytometry [20]. The sensitivity (95% confidence interval) is 99.2% (97.7–99.8) and the specificity is 100% (98.5–100) [21]. The assay is standardized with WHO international reference sera (20/136 and 20/130) and cross-validated with two commercially available ELISA (Abbott 147 and Beckmann 56) to allow calculation of BAU/mL [22]. Neutralizing activities in sera were measured with the S-Fuse assay using authentic isolates of Delta and Omicron. This assay uses U2OS-ACE2 GFP1-10 and GFP 11 reporter cells, also termed S-Fuse cells, that become GFP+ upon infection with SARS-CoV-2 [23,24]. All sera were tested in limiting dilution to determine Effective Dilution 50% (ED50 or titer) values. In each well, the number of GFP+ syncytia was scored with an Opera Phenix high-content confocal microscope (PerkinElmer). ED50 were calculated with a reconstructed curve using the percentage of the neutralization at the different concentrations. Viral stock were produced on Vero E6 cells, titrated on Vero E6 or S-Fuse cells and sequenced to confirm viral lineages (GISAID accession ID: EPI_ISL_2029113 and EPI_ISL_7413964 for Delta and Omicron isolates, respectively).

We also evaluated the risk of breakthrough infection among residents from two out of the three nursing homes where cases were recorded during the French Omicron epidemic wave in December 2021-January 2022. During this time, residents living in nursing homes where COVID-19 positive cases were reported were submitted to RT-PCR screening on at least two occasions, independently of the presence of symptoms. RT-PCR positive test results were reanalysed with a second round of RT-PCR screening to identify SARS-CoV-2 variants of concern based on mutations E484K, E484Q and L452R, or K417N.

### Ethical considerations

This study was registered with ClinicalTrials.gov (NCT04644159) and received ethical approval by the Comité de Protection des Personnes Nord Ouest IV. Informed consent was obtained from the residents, or their relatives when the residents did not have full capacity to sign legal documents.

## Results

Among the 38 elderly (30 females and 8 males) included in the study, with median (IQR) age of 88 (81-92) years at the time of last sampling, 30 (78.9%) had been previously infected, based on past clinical history and serological findings. Most infections (90.0%) took place in February-March 2020, including three individuals whose infection was diagnosed by positive serology in May 2020 in the absence of clinical symptoms or positive RT-PCR findings. For two other convalescent residents, the infection took place before the primary series of vaccination, and for one resident the time of infection could not be determined (detection of anti-N antibodies in sera sampled post-primary series, without prior samples available).

Following the second dose of vaccine, anti-Spike IgG levels were higher among convalescents compared with naive residents (Table 1 & Fig 1b). The third dose increased the antibody titers for naive residents to levels similar to those of the convalescents after 2 doses (Table 1 & Fig 1b). The ED50 of neutralization were lower against Omicron compared to Delta, and higher among convalescents compared to naive residents (Table 1 & Figure 1c). For the eight naive residents, neutralization was detectable for only five (62.5%) and one (12.5%) individuals against Delta and Omicron after the second dose, respectively. The number of residents displaying neutralizing antibodies increased to eight (100%) against Delta and six (75%) against Omicron after the third dose. All convalescents except one neutralized Delta and Omicron after the second and third dose.

**Table 1.** Median (IQR) anti-Spike IgG and ED50 of neutralization against Delta and Omicron. P values A vs D, B vs E, C vs F : Mann-Whitney test ; B vs C, E vs F : Wilcoxon test

|  | Naive |  |  | Convalescent |  |  | P values |  |  |  |  |
| --- | --- | --- | --- | --- | --- | --- | --- | --- | --- | --- | --- |
|  | anti-S IgG | ED50 Delta | ED50 Omicron | anti-S IgG | ED50 Delta | ED50 Omicron | A vs D | B vs E | C vs F | B vs C | E vs F |
|  | A | B | C | D | E | F |  |  |  |  |  |
| 2nd dose | 456<br>(210-571) | 37<br>(15-78) | 15<br>(15-15) | 3056<br>(1838-4604) | 12393<br>(3842-19571) | 1113<br>(256-2930) | 0.0001 | <0.0001 | <0.0001 | 0.06 | <0.0001 |
| 3rd dose | 1256<br>(1020-2747) | 1243<br>(473-4575) | 188<br>(51-787) | 2485<br>(1407-3271) | 6156<br>(2747-11919) | 1088<br>(477-2556) | 0.33 | 0.03 | 0.02 | 0.01 | <0.0001 |

In December 2021-January 2022, two out of the three nursing homes were affected by an Omicron epidemic, as reavealed by routine PCR testing (see Methods and Fig 2a). This epidemic took place three months after administration of the third dose. Among the 32 residents (27 females and 5 males) living in these two households, 12 breakthrough infections occurred (34%). Seventy-five percent (6/8) of naive residents were infected, compared to 25% (6/24) of convalescent (P=0.03; two-sided Fisher exact test). Anti-Spike IgG and ED50 of neutralization against Omicron post 3^rd^ dose were compared between those with and without breakthrough infection (corresponding to a median (IQR) of 53 (49-60) days before the breakthrough infection) (Fig 2b). Median titers were lower in those with compared to those without breakthrough infection : 1256 vs 2523 BAU/mL (P=0.016) for anti-Spike IgG; and 234 vs 1298 (P=0.0004) for ED50 against Omicron, respectively. Of note, none of the individuals with a ED50 titer above 1542 had breakthrough infection. The breakthrough infections were considered mild-to-moderate by the physicians in charge, requiring no hospitalization or oxygenotherapy.

**Figure 2:**
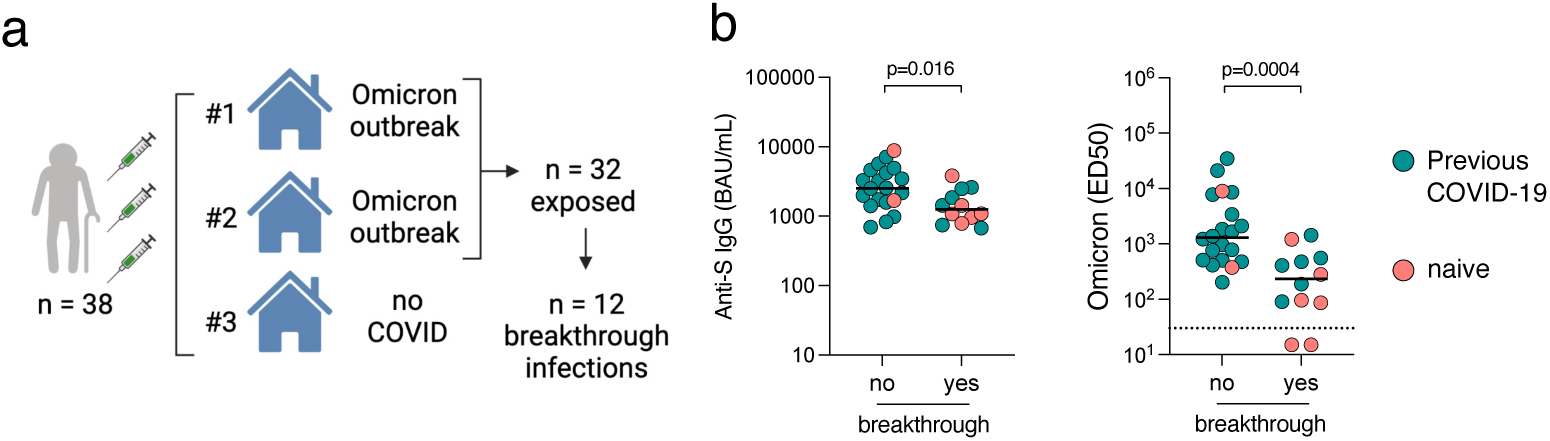
Humoral immune response predicts odds of Omicron breakthrough infection. (a) Omicron circulated in 2 out of the 3 nursing homes included in the study. In consequence, 32 individuals were exposed to the virus (27 females and 5 males) and 12 breakthrough infections occurred. (b) Levels of anti-Spike IgGs and neutralization of Omicron are indicated for individuals having a subsequent confirmed breakthrough infection or not. Data are provided as Binding Arbitrary Unit per mL (BAU/mL) (left) and neutralization titers (ED50) (right). Comparisons were performed using the Mann-Whitney rank test. Green dots indicate individuals with an history of COVID-19 prior to their booster dose of vaccine. Pink dots indicate individuals with no previous COVID-19. The dashed line indicates the limit of detection.

## Discussion

This study shows lower levels of anti-Spike IgG and neutralizing antibodies against Delta and Omicron among naive compared to convalescent nursing home residents after two doses of mRNA vaccine. A third dose of BNT162b2 vaccine significantly increased antibody levels for naive residents, showing the importance of receiving at least three antigenic exposures to the Spike protein for achieving satisfactory neutralizing antibody levels [25]. Nevertheless, median antibody levels and neutralization titers in naive residents remained lower than in convalescents, and were not sufficient to prevent infection with Omicron for most of them. This was particularly the case for those with the lowest levels of neutralizing antibodies, suggesting that a fourth antigenic exposure may be useful in this particularly aged population to prevent infection with Omicron, a variant known for its high escape immunity properties. The absence of severe forms of disease in the Omicron-infected group is reassuring, however based on a small sample size.

Anti-Spike IgG and neutralization titers were lower among residents who had breakthough infections compared to those who had not, regardless of COVID-19 history. No antibody level would however be discriminative enough to separate the two groups and be used as a correlate of protection. This may have to do in part with the very high escape immunity properties of the Omicron variant.

Most previous infections occured during the first epidemic wave (February-March 2020), so that the vaccine primary series performed in early 2021 was able to boost the production of neutralizing antibodies primed one year earlier, even in this elderly population. This is comforting, and reflects the strength of the so-called hybrid immunity combining the effects of infection and vaccination [26]. Interestingly, among these vaccinated-convalescent residents, antibody levels were slightly lower in samples collected two months after the third dose than in samples collected two months after the second dose. This may be explained by the waning of immune responses and low levels of circulating antibodies before the booster dose, so that the levels achieved after the third dose did not reach those achieved just after one infection and two doses.

Our study has limitations. First, the small sample size limits our statistical power and precludes analysis of the characteristics that may further impact vaccine efficacy, such as gender, prexisting conditions or ongoing medications. Second, we did not have access to nasopharyngeal swabs to measure antibody levels at the site of viral entry and replication. We were thus unable to link breakthrough infections to local levels of antibodies, which might represent a better correlate of protection. Further studies are needed to determine the contribution of mucosal immunity on the acquisition of SARS-CoV-2 and the severity of COVID-19. Lastly, we only tested BA.1, the initial Omicron clade, which was circulating in France at the time of the investigation and was responsible of the breaktrough infections that occurred in the nursing homes of the study. It will be worth examining the neutralization activity of the sera against other Omicron sub-lineages, such as BA.2, at the next blood sampling of the same cohort participants.

This study confirmed the importance of receiving at least three antigenic exposures to the Spike protein for achieving satisfactory neutralizing antibody levels against Omicron. Protection against Omicron was increased in those who had been previously infected in addition to the three doses, suggesting that a fourth antigenic exposure may be useful in this elderly population to prevent infection with a variant known for its high escape immunity properties.

## Data Availability

All data produced in the present study are available upon reasonable request to the authors

## Acknowledgments

We thank the nursing home residents who agreed to participate into the study and the medical teams who were involved in sample and data collection. We thank the teams of Crépy-en-Valois town hall and the Director and technical services of the local hospital for their help in the implementation of the COVID-Oise study. We thank the ICAReB technical team for management and distribution of the samples. We thank members of the Virus and Immunity Unit for discussions and help, Nathalie Aulner and the UtechS Photonic BioImaging (UPBI) core facility (Institut Pasteur), a member of the France BioImaging network, for image acquisition and analysis. The Opera system was co-funded by Institut Pasteur and the Région ile de France (DIM1Health).

## Authors contribution

Conceptualization and Methodology: TB, LP, DP, OS, AF

Cohort management and sample collection: LP, LT, CD, AK, SFP, LPdF, ER, NJ, MNU

Serological and seroneutralisation assays: TB, DP, IS, FP, FGB, MA, SP, TW, SC, SvdW, MW

Data assembly and manuscript writing: TB, LP, DP, LT, OS, AF

Funding acquisition: OS, AF Supervision: OS, AF

## Conflicts of interest

The authors have no competing interest to declare.

## Fundings

Work in OS lab is funded by Institut Pasteur, Urgence COVID-19 Fundraising Campaign of Institut Pasteur, Fondation pour la Recherche Médicale (FRM), the European Health Emergency Preparedness and Response Authority (HERA), ANRS-MIE, the Vaccine Research Institute (ANR-10-LABX-77), Labex IBEID (ANR-10-LABX-62-IBEID), ANR/FRM Flash Covid PROTEO-SARS-CoV-2, ANR Coronamito, and IDISCOVR, “TIMTAMDEN” ANR-14-CE14-0029, “CHIKV-Viro-Immuno” ANR-14-CE14-0015-01 and the Gilead HIV cure program. AF lab is funded by the INCEPTION project (PIA/ANR-16-CONV-0005) and the Labex IBEID (ANR-10-LABX-62-IBEID). The COVID-Oise cohort is funded by “Alliance Tous Unis contre le virus” Institut Pasteur, AP-HP and Fondation de France.

## Data availability

All data produced in the present study are available upon reasonable request to the authors

## Notes

### Competing Interest Statement

The authors have declared no competing interest.

### Author Declarations

This study was registered with ClinicalTrials.gov (NCT04644159) and received ethical approval by the Comite de Protection des Personnes Nord Ouest IV. Informed consent was obtained from the residents, or their relatives when the residents did not have full capacity to sign legal documents.

